# The Chicago River Reversal of 1900: Separating Myth from Measured Impact on Public Health

**DOI:** 10.1101/2025.10.14.25337993

**Authors:** Stephenson Strobel

## Abstract

The reversal of the Chicago River in 1900 is often credited with dramatically lowering mortality by eliminating sewage from the city’s drinking water. Contemporary accounts and historians have described it as ending Chicago’s typhoid crisis. Using ward-level mortality records and synthetic control methods, I find the immediate effects were more modest than claimed. The reversal reduced mortality by 4% (approximately 985 lives) in 1900, with effects concentrated in summer months consistent with reductions in waterborne disease. These results suggest Chicago’s dramatic mortality decline required complementary interventions to produce the dramatic reductions in mortality that were observed in the 1900s. This iconic case highlights that public health progress depends on layered measures rather than single “silver bullet” solutions.

## 1 Introduction

The reversal of the Chicago River in 1900 is often celebrated as one of the great triumphs of American public health. By engineering the river to flow away from Lake Michigan rather than into it, Chicago eliminated the direct discharge of sewage into its own drinking water supply. Contemporary observers and later historians have described the reversal as a decisive turning point that freed the city from routine epidemics of typhoid fever and other water borne illness.

Yet despite its iconic status, the actual impact of the reversal on mortality has rarely been evaluated with data. Most accounts assume that the project immediately and dramatically reduced deaths in Chicago, but this claim is more based on narrative than evidence. Contemporary newspaper accounts proclaimed the reversal would “free the city from the growing menace of contaminated water supply” [1]. Later descriptive histories similarly credited the reversal with ending Chicago’s typhoid crisis [2, 3, 4, 5]. Using modern causal methods, can we see whether mortality in Chicago fell sharply after January 1900 in keeping with the mythology of the reversal?

This paper re-examines the event using ward and district-level mortality data from Chicago, Baltimore, and Cincinnati in 1899 and 1900. Applying synthetic control methods, I find that the reversal did reduce mortality in Chicago, but the decline was modest and reduced annual mortality by 0.6 deaths per 1,000 population, or roughly 985 lives saved in 1900. These effects were concentrated in the summer months, consistent with reductions in waterborne disease. However, they fall short of the dramatic improvements often attributed to the river reversal.

This finding does not diminish the reversal’s importance, but rather contextualizes it within Chicago’s broader mortality transition. Revisiting this episode highlights the limits of single interventions in producing broad population health gains. Chicago’s large mortality declines in the early twentieth century required not only sewage diversion, but also complementary measures such as chlorination, water filtration, and broader sanitary reforms. Understanding the true scale of the river reversal’s effect underscores an enduring lesson in public health: progress is rarely the product of one engineering feat, but rather of cumulative, layered interventions over time.

## 2 Historical Background

In the 19th century American cities suffered from high mortality due to infectious diseases. It is estimated that anywhere from 21 to 42% of Americans had contracted typhoid at some point in their lives [6]. The epidemiological transition in the US has been in large part, attributed to the reduction of these types of diseases which resulted in wide-spread economic and health improvement [7]. Chicago was no exception to this rule and and often averaged a typhoid mortality rate of 0.73 per 1000 population before 1893 with extreme years having 1 to 1.5 deaths per 1000 population [8]. Chicago’s own epidemiological transition as well as that of the United States is in part attributed to public health interventions that helped clean the countries drinking water [9, 8, 10, 11].

During the 1800s, Chicago had become a city defined as much by its industrial growth as by its sanitary challenges which were interconnected. Incorporated in 1833 with barely 300 residents, it expanded rapidly as railroads, canals, and slaughterhouses drew workers and capital. The Union Stock Yards, opened in 1865, became the heart of Chicago’s meatpacking empire. Its waste was channeled into the South Fork of the Chicago River, via “Bubbly Creek” nicknamed for the gases rising from decomposing animal remains. By 1900, hundreds of millions of animals were slaughtered annually, and the refuse joined human sewage in flowing toward Lake Michigan where Chicago’s drinking water intake pipes were located [5].

The city’s leaders were aware of the danger. As early as the 1860s, engineers built intake tunnels further and further into Lake Michigan in an effort to draw cleaner water. But population growth and industrial expansion outpaced these measures. By the 1880s, Chicago’s mortality rate from typhoid fever was among the highest in the United States. The Chicago river itself had become an open sewer [1, 12] and fears of catastrophic epidemics were widespread. In 1877, heavy rain caused the river to empty sewage into Lake Michigan and turned the city drinking water “wine-colored” [13]. In 1885, a major storm drove a visible plume of sewage into the lake, alarming residents, even though no outbreak was definitively linked to the event [14].

Public concern culminated in 1889 when the Illinois legislature authorized the creation of the Sanitary District of Chicago [13]. Its mandate was ambitious: to construct a canal that would reverse the flow of the Chicago River, sending sewage westward toward the Des Plaines and Mississippi Rivers rather than eastward into Lake Michigan [14]. The project became one of the largest earth-moving efforts in North America before the Panama Canal, employing more than 15,000 workers at its peak [5].

Completed in 1899 and officially opened in January 1900, the Chicago Sanitary and Ship Canal was celebrated as a civic triumph. Civic leaders presented the reversal not only as an engineering victory but as the long-awaited solution to Chicago’s typhoid problem. Newspapers hailed it as a modern marvel that would permanently safeguard the city’s health. The Chicago Tribune described the city as “free from the growing menace of contaminated water supply” [1]. A letter to the editor published in JAMA described Chicago as having a “very serious question of how properly to dispose of its sewage” concluding that the river’s reversal “solves the whole problem” [15]. However, these proclamations were soon tempered. A typhoid outbreak in 1902 surprised Chicagoans [16] with the Chicago Tribune confessing that while without the canal “the water would be…rank poison” the “entire water supply is of inferior quality” [1]. This raises the question: how much did the reversal actually reduce mortality in its immediate aftermath?

The river reversal was preceeded and followed by additional interventions but the opening of the canal was relatively isolated from these. Water intake cribs were constructed far from the polluted shoreline were completed in in 1869. Sewage diversion occurred in 1907 and prevented any further dumping of refuse directly into Lake Michigan. Municipal milk purification occurred in 1909. Finally, chlorination of the city’s water supply began in 1917. Understanding the reversal’s isolated effect helps parse the contributions of these complementary measures [8, 17].

## 3 Methods

### 3.1 Data

I use the Historical Urban Ecological (HUE) dataset, which compiles monthly mortality rates for major US cities from the 19th and early 20th centuries [18]. For this study, I use ward and district-level data from Chicago, Baltimore, and Cincinnati covering January 1899 through December 1900. These three cities are the only ones in the dataset with monthly, ward-level coverage for this period. In total, the data include 34 wards in Chicago, 30 districts in Cincinnati, and 22 districts in Baltimore. Ward level population data is taken from 1896 for Baltimore and Chicago and for 1890 for Cincinnati which are the closest available years with ward level data.

The outcome of interest is the overall mortality rate, measured as deaths per 1,000 residents per month. Unfortunately, cause-specific mortality data (e.g., typhoid or diarrheal disease) are not available for Chicago in these years, which limits the ability to directly trace effects to waterborne illness. The short observation window is another limitation: Chicago reorganized its wards in 1901, preventing consistent ward-level analysis beyond the first year of the canal’s operation.

### 3.2 Estimation and Inference

To estimate the impact of the Chicago River reversal, I compared mortality in Chicago before and after January 1900 with mortality in Baltimore and Cincinnati. The districts in these cities serve as controls because they had similar urban structures and mortality profiles during this period but did not experience a comparable water infrastructure shock. The full econometric specification is found in Appendix A.

To isolate the reversal’s causal effect, I employ a comparative framework using Baltimore and Cincinnati as controls. These cities had similar characteristics to Chicago such as industrial economies, comparable population sizes, and similar baseline mortality rates. However these donor cities did not reverse their rivers or experience comparable water infrastructure shocks in 1899-1900.

I used two complementary approaches. First, a simple two-way fixed effects difference-in-differences (TWFE-DiD) framework compares mortality changes in Chicago wards before and after the reversal to changes in wards from the control cities over the same period. This design essentially takes all wards in Chicago and compares them to all wards in Cincinnati and Baltimore. Second, a synthetic control method constructs a weighted “synthetic Chicago” out of the ward-level data from Baltimore and Cincinnati. This is designed to closely match Chicago’s pre-reversal mortality trends. Divergence between Chicago and its synthetic counterpart after January 1900 provides an estimate of the river reversal’s effect.

To assess whether Chicago’s mortality decline could have occurred by chance, I conduct placebo tests. I randomly assign “treatment” to wards in Baltimore and Cincinnati and estimate pseudo-effects. If Chicago’s effect is unusually large relative to this placebo distribution, it suggests a genuine treatment effect rather than random variation.

## 4 Results

Figure 1 presents overall mortality trends and distributions for Chicago, Baltimore, and Cincinnati between January 1899 and December 1900. Mortality levels of approximately 1–2 deaths per 1,000 population per month (12–24 deaths per 1,000 per year) are consistent with early twentieth-century urban health patterns. The time series (1a) provides initial visual evidence: Chicago’s mortality did not surge in summer/fall 1900 as sharply as Baltimore and Cincinnati. This divergence is consistent with the reversal preventing waterborne disease deaths that would have occurred absent the intervention. Both subfigures confirm strong overlap between treated and control wards, supporting the support assumption underlying the research design.

**Figure 1:**
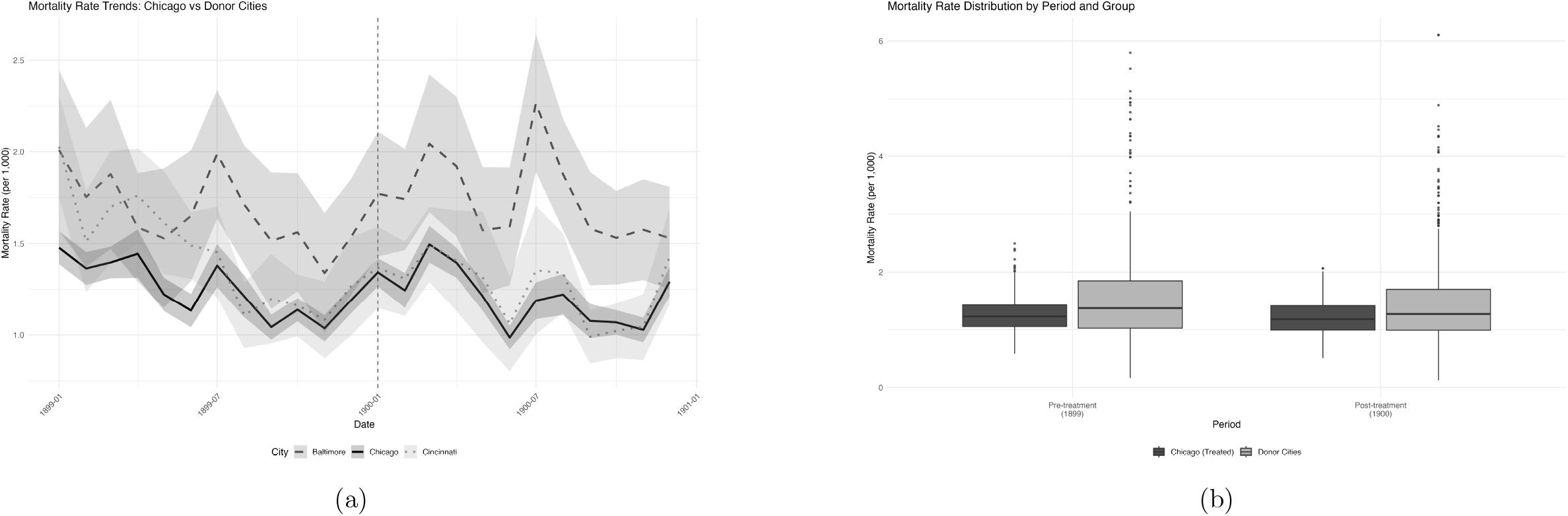
Mortality outcomes: (a) trends, and (b) distributions by donor vs. treatment. Chicago wards are compared to Cincinnati and Baltimore districts. There is good overlap between treatment and donor wards both over the year of observation and overall.

Figure 2a shows baseline mortality rates of Chicago wards and demonstrates elevated mortality in the downtown and northern wards. Figure 2b also demonstrates good fit by *R*^2^ measures between treatment wards and their synthetic controls during the pretreatment period.

**Figure 2:**
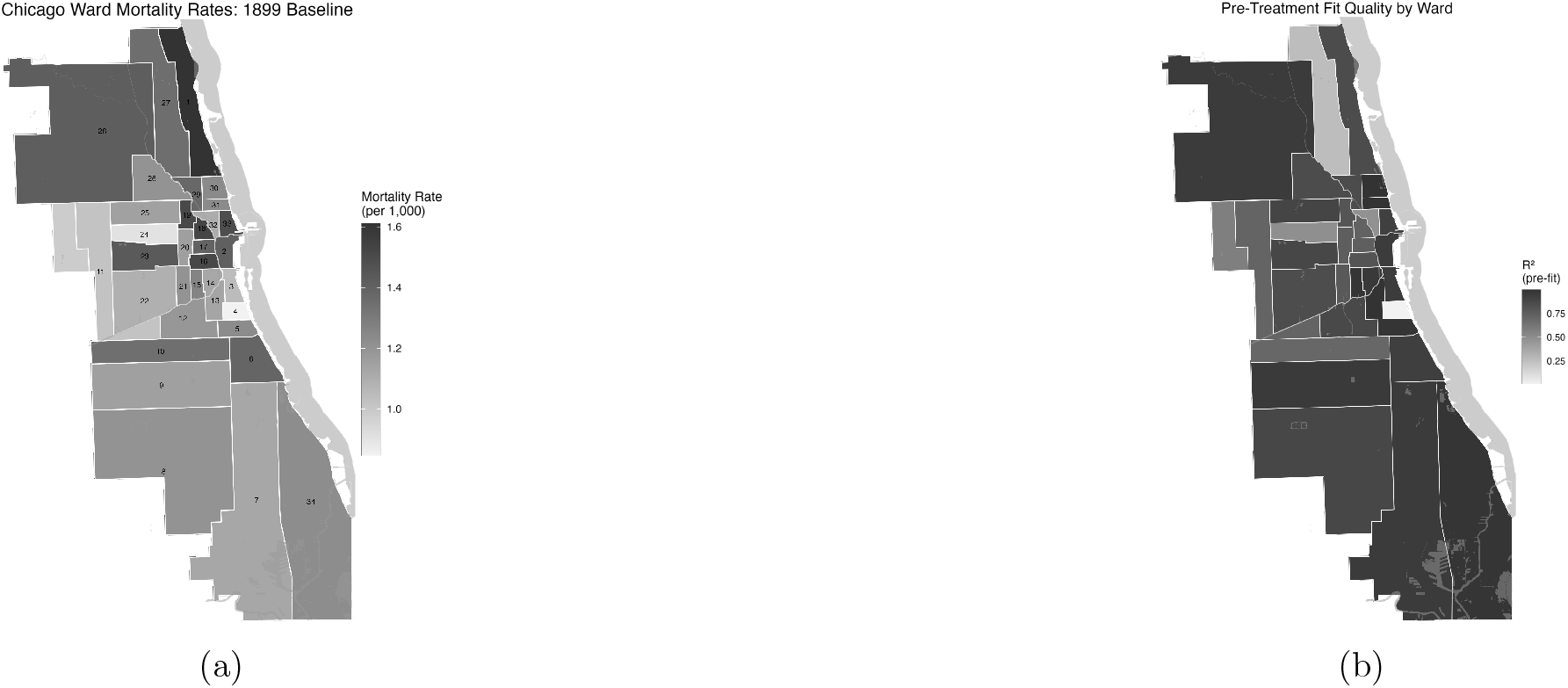
Geographic results: (a) Pre-treatment mortality and (b) mapped *R*^2^ fit. Pre-treatment mortality in Chicago is calculated as the average mortality across all months in 1899 for each ward. It is generally highest in the downtown area and in the north of the city along Lake Michigan. Pre-treatment fit of Chicago wards to donor synthetic wards is high as measured by *R*^2^.

Figure 3 plots Chicago ward effects against placebo effects from donor wards. Overall, effects are not statistically distinguishable from placebos, but when stratified by season, a clear pattern emerges: in summer and fall 1900, most Chicago wards (82%) show negative effects, while their pre-reversal distributions were centered at zero. This seasonal pattern is consistent with reductions in waterborne disease.

**Figure 3:**
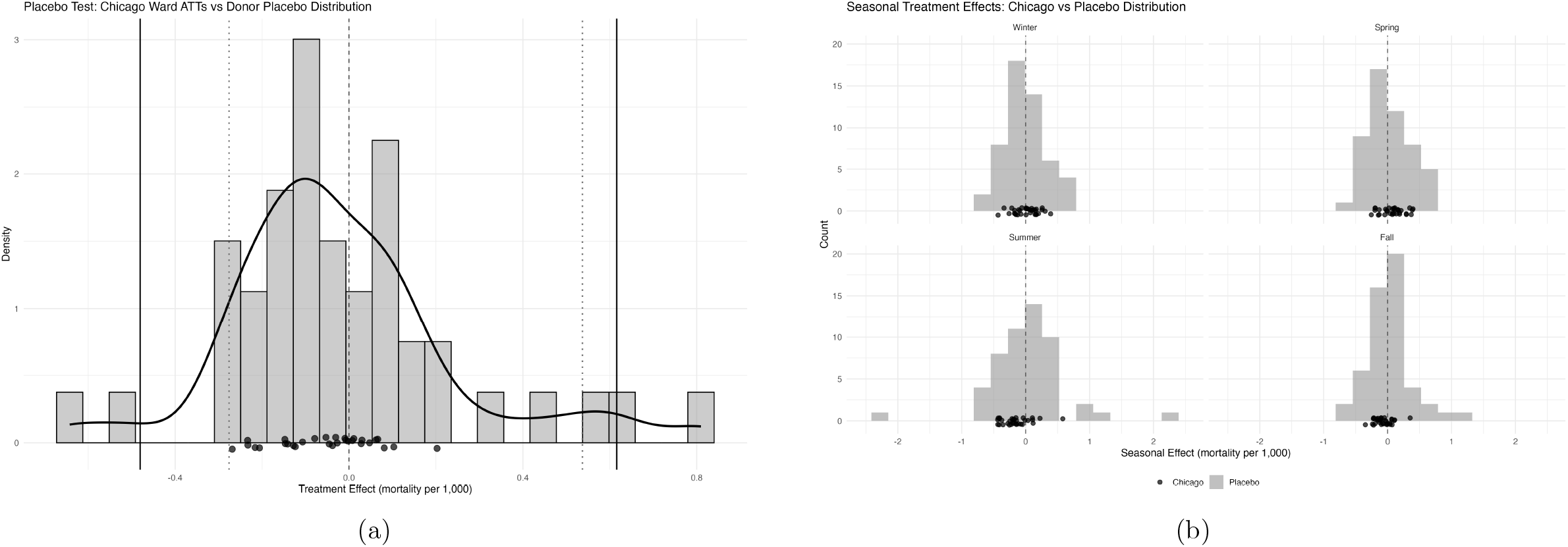
Comparison of placebo distributions: (a) overall, and (b) stratified by season. Across overall and season-based placebo distributions there are no statistically significant changes in mortality. However, in summer and fall months, the distribution of effects on treatment wards in Chicago shifts from an equal distribution around zero to predominantly negative in magnitude.

A first set of causal estimates using a TWFE event study is shown in Figure 4a. In July 1900, mortality in treated Chicago wards declined by 0.36 deaths per 1,000 population relative to Baltimore and Cincinnati. This pattern is consistent with reductions in waterborne disease. However, pre-treatment trends suggest that the parallel trends assumption may be violated, raising concerns about the validity of these estimates. The overall TWFE-DiD effect suggests an increase in mortality of 0.03 deaths per 1000 population, although this is not significantly different than zero.

**Figure 4:**
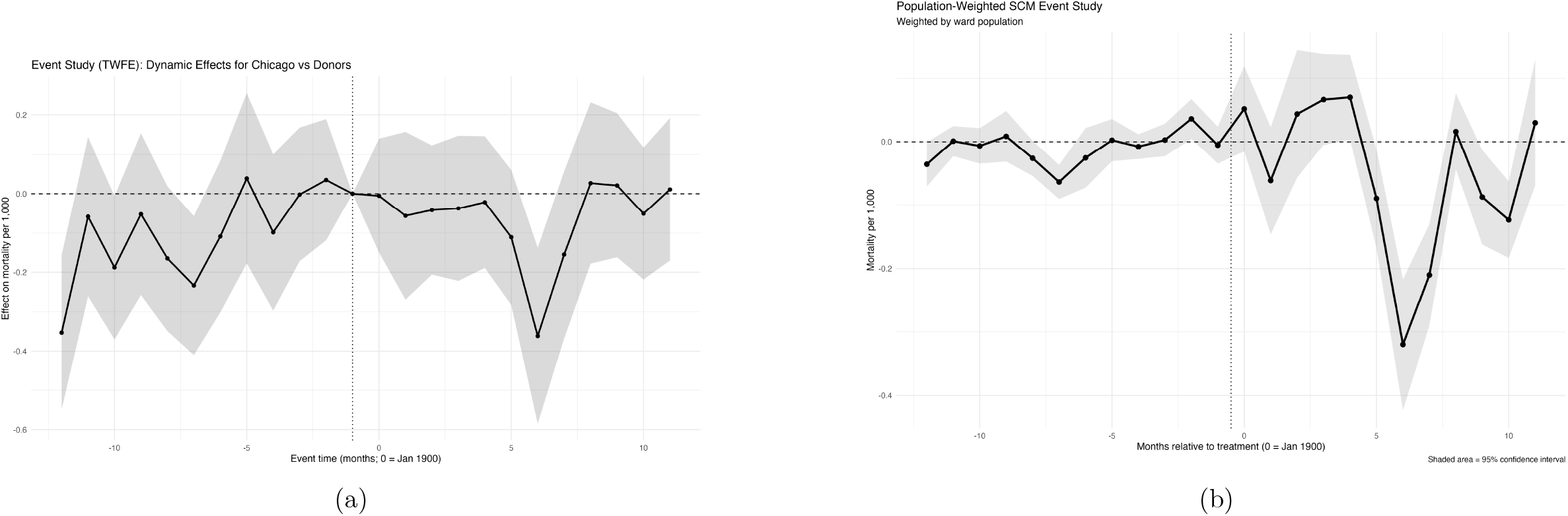
Event Studies: (a) TWFE Event Study, and (b) Population weighted average synthetic control method effects. TWFE events demonstrates significant improvement in July 1900 with some evidence of violation of the pre-trends assumption. SCM events suggest similar effects in summer months and later in the fall. There is much less evidence of pre-trends violation and effects are more precisely estimated.

The TWFE pre-treatment trends violations motivate the synthetic control approach, which explicitly matches pre-treatment mortality trajectories rather than assuming parallel counterfactual trends. Figure 4b displays results from a population-weighted synthetic control event study. As with the TWFE analysis, I find sharp reductions in mortality during the summer months of 1900. In contrast to TWFE, these synthetic control results suggest more consistent declines throughout the summer and fall, and the pre-treatment fit better satisfies the assumption of parallel trends.

At the ward level (Figure 5), estimated effects range from −0.269 deaths per 1000 population per month in ward 8 to 0.202 per 1000 population per month in ward 21. Seasonal differences are evident: effects in the spring range from −0.431 to 0.392 deaths per 1000 population per month with 38% of wards displaying beneficial effects. Summer effects range from −0.437 to 0.580 with 82% of wards displaying beneficial effects. This shift aligns with reduced waterborne disease transmission in Chicago during warmer months.

**Figure 5:**
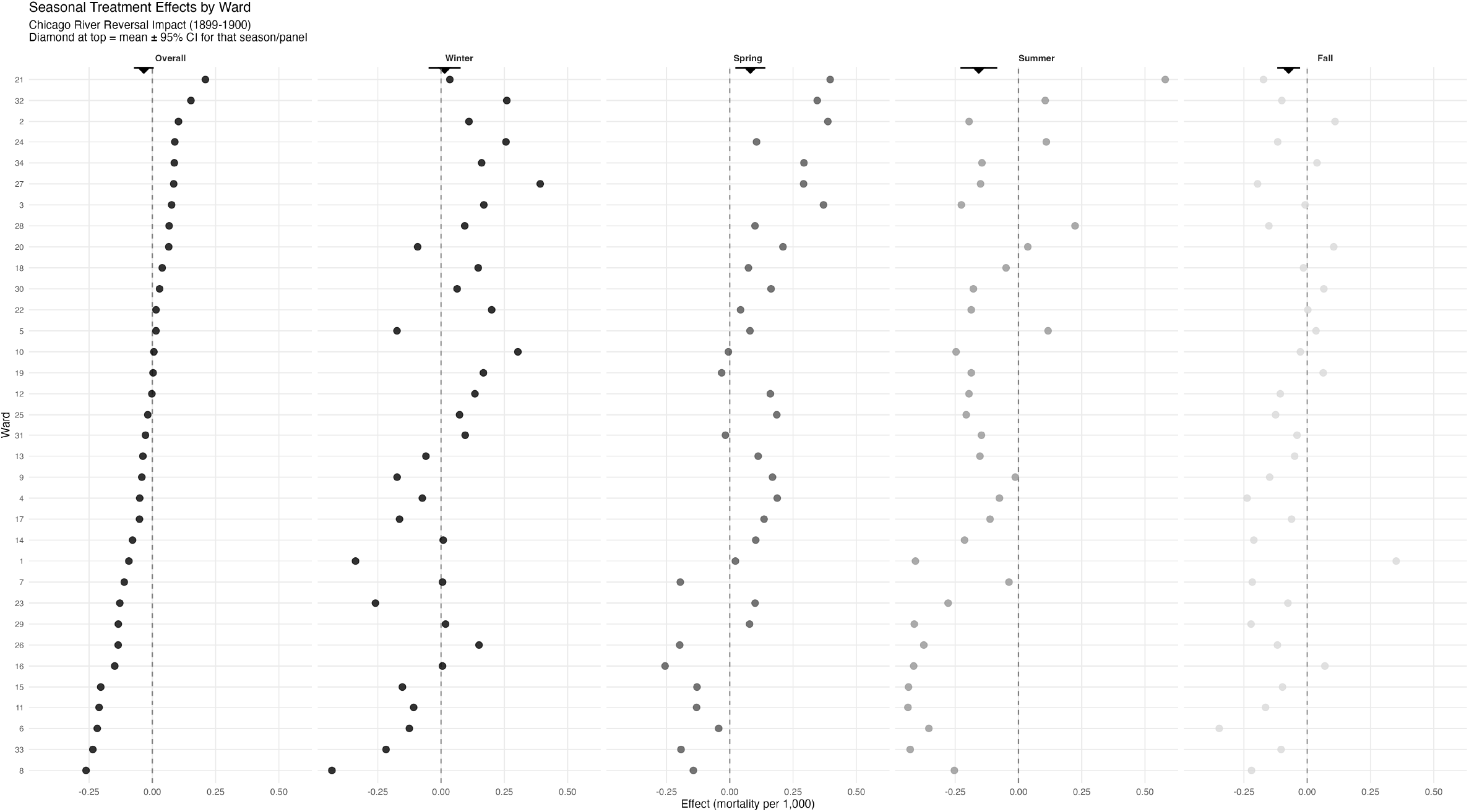
Effects of the reversal by ward and season. Individual SCM effects are plotted by ward with aggregated effects at the top of each seasonal plot with 95% CI.

Aggregating across all wards and months in 1900, the reversal is estimated to have reduced mortality by 0.61 deaths per 1,000 population per year. This corresponds to the prevention of approximately 985 deaths in a city of 1.6 million people. This is a reduction of 4.1% from the baseline mortality rate in Chicago.

## 5 Discussion

This study provides new evidence on one of the most celebrated public health interventions in American urban history. The reversal of the Chicago River in 1900 is often remembered as the decisive intervention that ended the city’s typhoid crisis by diverting sewage away from its drinking water supply. Using ward-level mortality data and modern synthetic control methods, I find that the immediate impact on mortality was real but modest. The reversal reduced mortality by approximately 0.61 deaths per 1,000 population annually in 1900 or roughly 985 lives saved in a city of 1.6 million residents. This represents a 4.1% reduction from baseline mortality, meaningful but far smaller than the dramatic transformation described in contemporary accounts and later histories.

Why did contemporaries overstate the reversal’s immediate impact? Several factors explain this disconnect. First, the absence of cause-specific mortality data made attribution difficult. The reversal was also highly visible and costly, creating political pressure to tout its success [1]. When mortality declined in summer 1900, the reversal received credit. When typhoid resurged in 1902, other factors were blamed [16]. This pattern of attributing successes to favored interventions while explaining away failures is common in public health history.

Why were the reversal’s immediate effects more modest than expected? Three factors help explain this finding. First, Chicago’s water intake pipes extended two miles into Lake Michigan by 1900 [14]. This distance provided substantial dilution even before the reversal, meaning the intake water was not as severely contaminated as the river itself. The reversal improved water quality at the margin rather than transforming it from sewage to pristine water.

Second, typhoid and other waterborne diseases, while significant, represented only a portion of Chicago’s overall mortality burden. In 1899, typhoid accounted for approximately 0.73 deaths per 1,000 population or roughly 1,240 deaths annually [8]. My estimate of 985 prevented deaths in 1900 suggests the reversal prevented a substantial share of waterborne disease mortality but could not address the many other causes of death prevalent in turn-of-the-century cities: tuberculosis, pneumonia, infant diarrhea from contaminated milk, and communicable diseases spread through crowded housing.

Third, the reversal addressed only one pathway of disease exposure. Contaminated milk remained a major source of typhoid and diarrheal disease until pasteurization ordinances passed in 1908. Person-to-person transmission continued in crowded tenements. Ground water contamination from privies and cesspools persisted in neighborhoods without sewer connections. The reversal was necessary but not sufficient for comprehensive mortality reduction.

The seasonal pattern of effects reinforces this interpretation. Reductions concentrated in summer and fall months, when 82% of wards showed beneficial effects compared to only 38% in spring. This timing aligns precisely with waterborne disease transmission patterns in this era, when warmer temperatures accelerated bacterial growth and residents relied more heavily on potentially contaminated water sources. However, the absence of statistically robust declines across all individual wards highlights the limits of attributing sweeping public health transformations to a single intervention.

My finding of a 4% mortality reduction may appear modest compared to previous literature on clean water interventions in the early 1900s. Cutler & Miller [9] conclude that clean water technologies accounted for nearly half of US mortality decline during this period. Ferrie & Troesken [8] find similarly large impacts on Chicago specifically from water purification. Phillips et al. [19] demonstrate that water supply investments were highly cost-effective, with a $1 per capita investment reducing typhoid transmission by 5%. Knutsson [20] finds that historical water purification in Sweden prevented significant mortality during a cholera outbreak.

However, these findings are complementary rather than contradictory. First, I examine the immediate, isolated effect of one specific intervention in a single year. Cutler and Miller [9] examine the cumulative effect of multiple technologies (filtration, chlorination, sewage treatment) implemented over several decades across many cities. The difference reflects timing and scope rather than inconsistent results.

Second, my one-year observation window cannot capture longer-run mortality effects. Reductions in waterborne diseases produce mortality declines beyond the immediate disease category through what early public health researchers called the Mills-Reincke phenomenon [21]. Preventing typhoid infection in childhood, for instance, may reduce vulnerability to other infections later in life by preserving health capital and avoiding chronic complications. My estimates capture only the immediate mortality effect, not these longer-run health improvements or cumulative benefits as cleaner water reduced the disease burden over time.

Third, recent work has refined our understanding of clean water’s effects. Anderson et al. [17] argue that clean water technology may have reduced child mortality sub-stantially but had smaller effects on adult mortality and total mortality than initially claimed. Their focus on average treatment effects across many cities and interventions differs from my single-intervention, single-city approach. Moreover, effects of individual interventions vary considerably across cities and may interact in complex ways [11]. What worked in one urban context may have been less effective in another, and interventions implemented simultaneously may have complementary or synergistic effects that exceed the sum of their individual contributions.

The modest immediate effect underscores the point that the reversal was a necessary first step but not a complete solution. Chicago’s dramatic mortality decline over the following decades required multiple, layered interventions. The city implemented comprehensive water filtration in 1917, began systematic chlorination in 1916, and passed milk pasteurization ordinances in 1908 [6]. By 1920, Chicago’s typhoid mortality rate had fallen to essentially zero [8].

Evidence from other geographies suggests these interventions had complementary effects. Alsan & Goldin [22] demonstrate that in Massachusetts cities between 1880-1920, water supply improvements and sewerage systems were most effective when implemented together rather than sequentially, with combined benefits exceeding the sum of individual effects. The river reversal likely enabled subsequent interventions to be more effective by reducing pathogen loads in source water, making filtration and chlorination more feasible and cost-effective. Filtration plants designed for moderately contaminated lake water would have been overwhelmed by direct sewage discharge.

The evidence for complementarity has important implications for understanding historical mortality decline and for contemporary policy. It suggests that cross-city studies estimating average effects of single interventions may understate their importance when implemented as part of comprehensive programs. An intervention that appears modestly effective in isolation may prove transformative when combined with complementary measures. Conversely, policymakers expecting dramatic results from single interventions may be disappointed if supporting infrastructure and practices are absent. This finding has implications for how we interpret the historical mortality transition. It suggests that Cutler & Miller’s [9] influential conclusion about clean water’s role should be understood as reflecting the cumulative effect of multiple technologies rather than any single intervention.

Most importantly, this historical case illustrates an enduring lesson for public health policy: progress rarely results from silver-bullet solutions. Transformative improvements emerge from sustained investment in multiple, complementary interventions that address disease transmission through various pathways. This lesson, learned in Chicago over decades, remains relevant for cities confronting water quality and sanitation challenges today. Over 2 billion people globally lack access to safely managed drinking water, with urban populations in developing countries facing conditions similar to 1900s Chicago. Understanding that single interventions are unlikely to transform outcomes alone can help set realistic expectations and sustain political commitment to comprehensive, long-term public health investment.

Recognizing the modest immediate effects of the river reversal does not diminish its historical importance. Rather, it contextualizes the intervention within Chicago’s broader mortality transition and highlights how public health progress depends on cumulative, multifaceted approaches sustained over time. This more nuanced understanding of a celebrated public health success enriches our appreciation of how cities overcame the sanitary crises of the industrial era and provides guidance for contemporary challenges.

## Data Availability

All data and code to replicate results in the present study are available upon reasonable request to the authors

## A Appendix A. Technical Methods

### A.1 Difference-in-Differences Estimation

I begin with a standard two-way fixed effects difference-in-differences specification [23]:

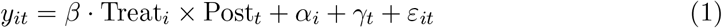

where *y*_*it*_ represents mortality per 1,000 in unit *i* at time *t, α*_*i*_ are unit fixed effects, *γ*_*t*_ are time fixed effects, and *ε*_*it*_ is the error term. The coefficient *β* captures the average treatment effect on the treated (ATT). I cluster standard errors at the ward level to account for serial correlation within units. To assess pre-treatment parallel trends and dynamic treatment effects, I also estimate an event study specification:

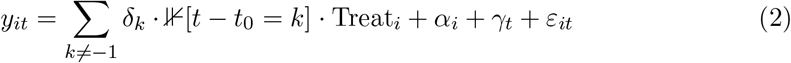

where *k* indexes event time relative to treatment (*t*_0_ = January 1900), and *k* = *−*1 (December 1899) serves as the reference period.

### A.2 Synthetic Control Method

While DiD provides an overall treatment effect, it relies on parallel trends assumptions which may not hold. To address these limitations, I implement ward-level synthetic control estimation following [24].

For each Chicago ward *j*, I construct a synthetic control as a weighted average of donor units that best reproduce the ward’s pre-treatment mortality trajectory. Let *Y*_1*t*_ denote the mortality per 1000 population for treated ward *j* at time *t*, and let *Y*_0*t*_ represent the (*N*_0_ *×* 1) vector of outcomes for the *N*_0_ donor units. The synthetic control weights 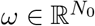 solve:

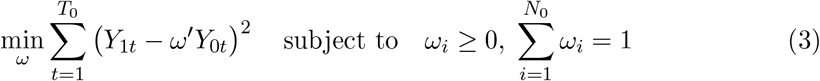

where *T*_0_ = 12 denotes the pre-treatment period. I estimate synthetic controls separately for each of the 34 Chicago wards, allowing donor weights to vary across treated units. The ward-specific treatment effect in period *t* is:

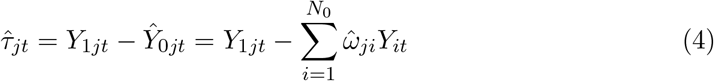

I evaluate pre-treatment fit quality using the coefficient of determination (*R*^2^) over the pre-period. For both TWE-DiD and SCM, the assumption is that the weighted combination of donor units provides a valid counterfactual for Chicago, which requires: (1) no anticipation of treatment, (2) no spillover effects onto control/donor units, and (3) the control matches Chicago’s pre-treatment trajectory.

### A.3 Aggregation to City-Level Effects

To obtain city-level treatment effects from ward-specific synthetic controls, I compute population-weighted averages. For each period *t*, the aggregate effect is:

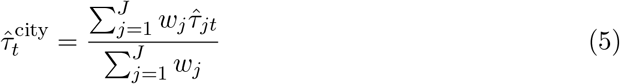

where *w*_*j*_ denotes ward *j*’s population and *J* = 34 is the number of Chicago wards. The standard error of the aggregate effect is computed as:

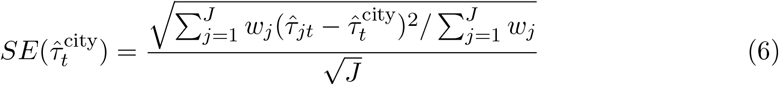

Confidence intervals are constructed as 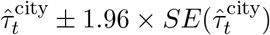.

### A.4 Inference via Placebo Tests

Following [25], I conduct in-space placebo tests to assess statistical significance for my SCM DiDs. I apply synthetic control procedures to each donor unit, treating it as if it received the intervention. This generates a distribution of placebo treatment effects under the null hypothesis of no effect. For each Chicago ward *j*, I compute a rank-based *p*-value:

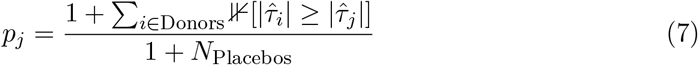

where 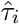 denotes the average post-treatment effect for placebo unit *i*.

## Notes

### Competing Interest Statement

The authors have declared no competing interest.

### Funding Statement

This study did not receive any funding

